# Effectiveness and Safety of Niclosamaide as Add-on Therapy to the Standard of Care Measures in COVID-19 Management: Randomized controlled clinical trial

**DOI:** 10.1101/2021.06.10.21258709

**Authors:** Ahmed S. Abdulamir, Faiq I. Gorial, Sattar Jabar Saadi, Mohammed Fauzi Maulood, Hashim Ali Hashim, Manal K. abdulrrazaq

## Abstract

**Background:** COVID-19 pandemic has ignited the urge for repurposing old drugs as candidate antiviral medicines to treat novel challenges of viral infections. Niclosamide (NCS) is an anti-parasitic drug of known antiviral potential. Therefore, this study attempts to investigate the antiviral effect and safety of NCS on SARS-CoV-2 caused COVID-19 patients.

**Methods:** Randomized controlled open label clinical trial encompassed 75 COVID-19 patients treated with standard of care plus NCS were included as experimental group and 75 COVID-19 patients treated with only standard of care therapy as control group. Each group was composed of 25 mild, 25 moderate and 25 severe patients. Survival rate, time to recovery, and side effects were the main endpoints for the assessment of the therapeutic effect and safety of NCS.

**Results:** NCS did not enhance survival rate as three of severe COVID-19 patients in NCS and in control groups died (P>0.05). However, NCS, compared to control group, reduced the time to recovery in moderate and severe COVID-19 patients about 5 and 3 days, respectively but not in mild patients (P≤0.05). Most interestingly, NCS lowered time to recovery up to five days in patients with co-morbidities (P≤0.05) whereas only one day lowered in patients without co-morbidities (P>0.05).

**Conclusion:** It is concluded that NCS accelerates time to recovery about 3 to 5 days in moderate to severe COVID-19 patients especially those with co-morbidities; hence, NCS is of clinical benefit for freeing hospital beds for more patients in pandemic crisis.

## Introduction

Coronavirus disease 2019 (COVID-19) is an infectious disease caused by severe acute respiratory syndrome coronavirus 2 (SARS-CoV-2) [1]. The outbreak of COVID-19 has become gradually nationwide with significant impact on public health and this outbreak was declared as a public health emergency on 30 January 2020 by the WHO and need an urgent viral infection identification and intervention as early as possible [2,3]. Several treatment strategies have been evaluated for Covid19 [4-6] but no current evidence from randomized controlled trials to recommend them as antiviral treatment for patients with COVID-19. On October 22, 2020 FDA approved NDA214787 for velkury (Remdesivir) which is indicated for adult and pediatric patients for treatment of COVID 19 requiring hospitalization [7] however WHO recommends against the use of remdesivir in COVID-19 patients [8].

Niclosamide was discovered in 1958. It is on the World Health Organization’s List of Essential Medicines. Niclosamide is an anti-parasitic medication used to treat tapeworm infestations. This includes diphyllobothriasis, hymenolepiasis, and taeniasis. In addition, Niclosamide may have broad clinical applications for the treatment of diseases other than those caused by parasites. These diseases and symptoms may include cancer, bacterial and viral infection, metabolic diseases such as Type II diabetes, NASH and NAFLD, artery constriction, endometriosis, neuropathic pain, rheumatoid arthritis, sclerodermatous graft-versus-host disease, and systemic sclerosis [9]. It is an FDA approved anthelmintic drug with low cost and low in vivo toxicity profile [10]. Recent drug repurpose screening identified NCS as an antimetabolite, antibacterial and anticancer agent [11,12]. Compelling body of evidences also suggest NCS possess broad spectrum antiviral properties including SARS-CoV (IC50 = 1.56 µM) [13,14].

Recently, it was also reported that NCS exhibited *in vitro* antiviral activity against SARS-CoV-2 (IC50 = 0.28 µM) [15]. When treating human subjects with a 2g oral dose of NCS, the maximum serum concentration was shown to be 0.25-6 ug/ml (0.76-18.3 uM). Hence, it is feasible to reach the effective IC50 of NCS on SARS-CoV-2 (0.28uM) by using the current dosage limits. However, the half-life of NCS is short, 6-7 hours [16]. Hence, to attain plasma concentrations of NCS close to the SARS-CoV-2 IC50 with less fluctuations, a daily oral dose of NCS 1g three times a day is needed.

The potential antiviral mechanism of NCS against SARS-CoV2 can be 1) blocking endocytosis of SARS-CoV2 [17] and 2) preventing autophagy of SARS-CoV-2 by inhibition of S-Phase kinase associated protein 2 (SKP2) [18]. Therefore; NCS can be a potential drug candidate for COVID-19 therapy.

This study was designed to assess the effectiveness and safety of Niclosamide as an addon therapy to the standard measures in COVID-19 management comapred to standards of care therapy group.

## Patients and Methods

### Study design

This pilot randomized controlled open label study was conducted at Alkarkh and Alforat hospitals in Baghdad city from January 2021-to April 2021. The study was performed according to the Declaration of Helsinki and its amendments, and the Guidelines for Good Clinical Practices issued by the Committee of Propriety Medicinal Product of the European Union. Ethical approval was taken from Iraqi Ministry of Health-Arab Board of Health Specialization in Iraq and the study was registered with the number of 20201541 at December 31, 2020T. Also, this study was registered in Clinical Trials.gov website under identifier number: NCT04753619. Informed consent was obtained from the participants to admit the study.

### Participants

#### Inclusion criteria

1. Patients with age above 18 years and of any gender.
2. Definite diagnosis of COVID-19 according to the WHO classification criteria [18].
3. Patients symptomatic for no more than three days for mild-moderate cases, no more than two days after being severe cases, and no more than one day after being critical cases.
4. Understands and agrees to comply with planned study procedures.

#### Exclusion criteria

1. Patients refuse to enrol in the study.
2. Patients with hypersensitivity or severe adverse effects to niclosamide.
3. Renal impairment (serum creatinine> 2 mg/dl).
4. Hepatic impairment(Alanine Aminotransferase (ALT) or aspartate aminotransferase (AST) > 3 times upper limit of normal.
5. Pregnancy or a desire to become pregnant.
6. Breast feeding.

In all the above condition Niclosamide should be avoided.

### Randomization and sample size calculation

This study is a randomized controlled trial with 2 arms study trial 1:1 allocation (75 Niclosamide group vs. 75 control group). Inside each group, up to 25 mild, 25 moderate and 25 severe and/or critical patients were recruited. The randomization process, patient’s records for disease progression, recovery, and clinical or laboratory testing were supervised by third party authority from Ministry of Health.

## Method

Patients were recruited from inpatients and outpatients according to the WHO classification criteria of severity of the disease. Study groups were divided into two groups:

NCS group: NCL + standard therapy.

Control group: Standard therapy used for mild-moderate or severe cases.

### Protocol of therapy

#### Niclosamaide Add-on group

– NCS 2 grams orally loading dose chewable then 1g every 12 hours were admisntered in the first day, then on the 2^nd^ day; 1g x3 for 7 days. That is to say, only in the first day 4 gm/d then on the second day 3g/d in 3 divided doses for 7 days.
– If the participant requires mechanical ventilation over the course of the study, NCS may be administered via nasogastric (NG) or orogastric (OG) tube and, if possible, should be administered with a scheduled nasogastric (NG) or orogastric (OG) feeding.

#### Control group

The patients in this group received only standard of care which included all or some of the following, according to the clinical condition of each patient:

– Acetaminophen 500mg on need.
– Vitamin C 1000mg twice/ day.
– Zinc 75-125 mg/day.
– 12 mg ivermectin single dose and 200 mg doxycycline on day 1, followed by 100 mg every 12 h for the next 4 days
– Favipravir course of therapy (1600mg twice daily in the first day then 600mg twice daily for up to 10 days total)
– Vitamin D3 5000IU/day.
– Azithromycin 250mg/day for 5 days.
– Oxygen therapy/ CBAP if needed
– Dexamethasone 6 mg/day or methylprednisolone 40mg twice per day, if needed.
– Mechanical ventilation, if needed.

Clinical, laboratory, and radiological evaluation used for diagnosis and evaluation of COVID-19 patietns;

1. Clinical evaluation: Gender, weight, height, smoking status. History of fever, dry cough, chest tightness or pain, SOB, sore throat, loss of taste, loss of smell, flulike symptoms, history of comorbid diseases, history of taking immune suppresseive drugs like streoids, immune suppressive agents, and biological therapy.
2. Laboratory investigations: Complete blood count (CBC), erythrocyte sedimentation rate (ESR), C-reactive protein (CRP), LDH, serum ferritin, d-dimer if present, liver function tests (ALT, AST), renal function tests (blood urea, serum creatinine), fasting blood sugar test, and nasopharyngeal swab, oropharyngeal swab for PCR.
3. Imaging evaluation:
  – Chest x-ray: non-specific imaging findings most commonly of atypical or organizing pneumonia, often with a bilateral, peripheral, and basal predominant distribution.
  – CT scan: suggestive of bilateral lung involvement most often as subpleural and peripheral areas of ground-glass opacity and consolidation.

### Outcome Measurements

A. Primary outcomes:
  – To assess the percentage of cure of the patient and evauated by normalization of clinical evaluation, laboratory investigations, and imaging.
  – To study the time to recovery.
B. Secondry outcomes:
  – Percentage of progression to more advanced disease.
  – Mortality rate among NCS add on group compared to controls.
  – Side effects seen during the trial which will be asseeded according to clinical evaluation and the appropriate laborartory investigation.

### Statistical analysis

Data was processed using SPSS version 13.1 and Microsoft Excel version 10.0.2. The quantitative data were subjected to normalization tests, normal distribution data were compared in terms of mean values to each other using student t-test while qualitative nominal data were analyzed using Chi square and Fisher Exact tests. P-value less than 0.05 is considered significant.

## Results

### Population characteristics

#### Participant flow

A total of 200 patients infected with COVID-19 were screened for elligiblity to be included in the study. Of those 50 patients were excluded: 2 of them were pregnant, 2 patients had elevated liver enzyme more than 3 times the normal reference range, one patient was <18 years age, 15 patients had heart failure, 10 patients had sepsis, 8 patients had liver failure, 7 patients had renal failure, 5 patients had rheumatoid arthritis on hydroxychloroquin amd salfasalazine. The remaining 150 patients were randomized to 75 Niclosamide add on group and 75 standards of care therapy group. All of the patients in both groups complteted the study as in figure1.

**Figure1:**
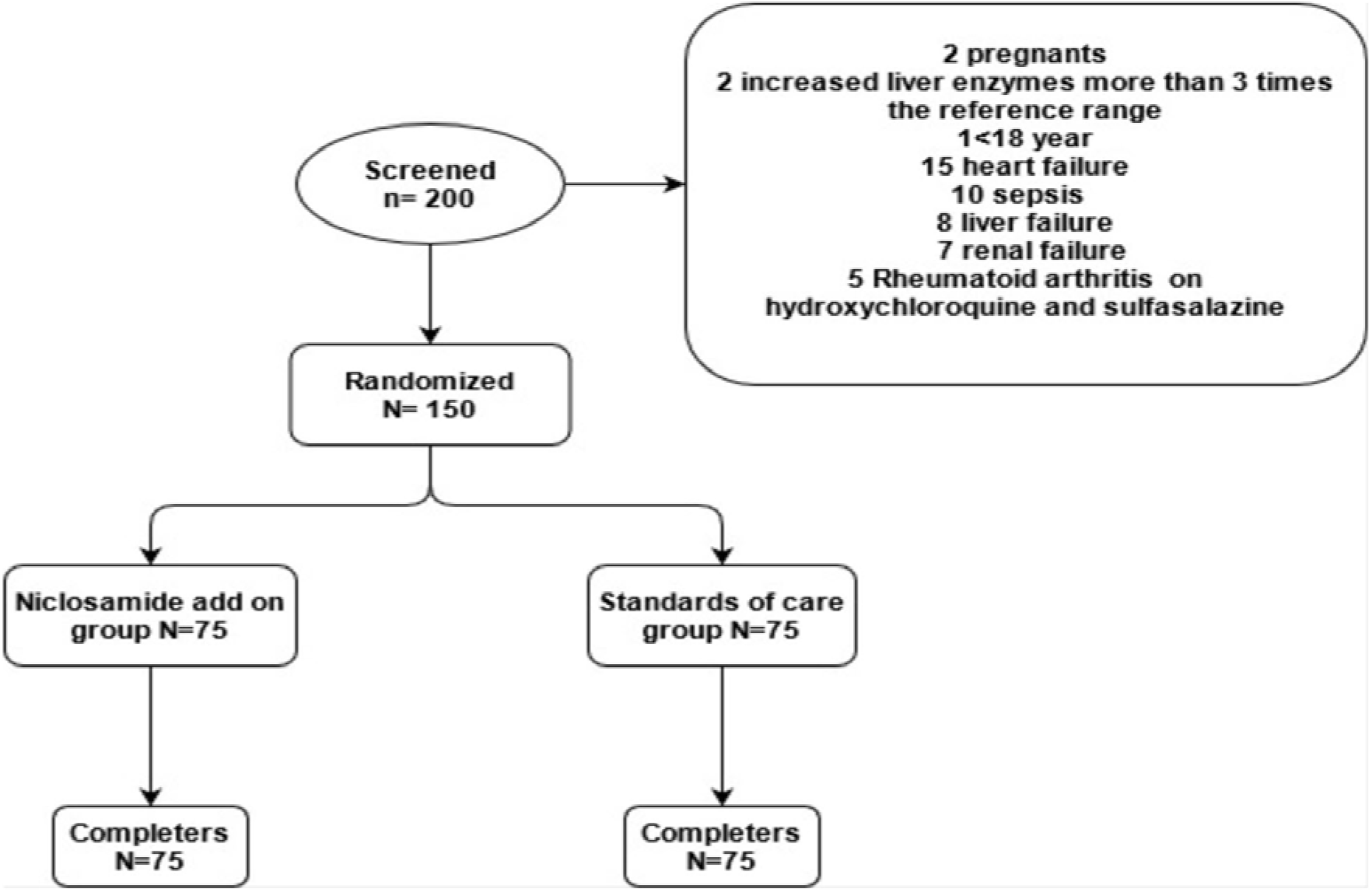
Study flow chart

### Baseline characteristics

The recruited patients were 70 females and 80 males; the average age of treated patients in NCS and control groups was, 49.29 (15.97) years and 49.32 (16.15 years, respectively (P=0.992). Minimum and maximum age in NCS and control groups were 16-82 and 15-95 years, respectively. The BMI of patients in NCS and control groups was 28.80 (5.35) kg/m^2^ and 30.01 (16.83) kg/m^2^, respectively (P= 0.556). Both arms of the study showed that COVID-19 patients’ mean presentation time was 7.81 (3.25) days in NCS and 7.11 (3.70) days in control group after onset of the disease (P=0.216) other baseline characteristics are shown in table1.

**Table 1:** Baseline characteristics of the niclosamaide add on group and controls

| Variables | Group | NCS add on group (n=75) | Controls (n=75) | p.value |
| --- | --- | --- | --- | --- |
| age mean (SD), years |  | 49.29 (15.97) | 49.32 (16.15) | 0.992 |
| sex n(%) | Female | 36 (48.0) | 34 (45.3) | 0.87 |
|  | Male | 39 (52.0) | 41 (54.7) |  |
| BMI mean (SD), kg/m<sup>2</sup> |  | 28.80 (5.35) | 30.01 (16.83) | 0.556 |
| smoking hx (%) | None smokers | 53 (70.7) | 56 (74.7) | 0.709 |
|  | Smokers | 22 (29.3) | 19 (25.3) |  |
| Time at presentation mean (SD), days |  | 7.81 (3.25) | 7.11 (3.70) | 0.216 |
| Presence of comorbidities n (%) | Absent | 34 (45.3) | 36 (48.0) | 0.87 |
|  | Present | 41 (54.7) | 39 (52.0) |  |
| comorbidities n (%) | Asthma | 1 ( 2.3) | 0 ( 0.0) | 0.394 |
|  | Obesity | 0 ( 0.0) | 1 ( 2.4) |  |
|  | HT | 13 (30.2) | 6 (14.3) |  |
|  | RA | 1 ( 2.3) | 0 ( 0.0) |  |
|  | IHD | 2 ( 4.7) | 0 ( 0.0) |  |
|  | DM | 3 ( 7.0) | 9 (21.4) |  |
|  | Epilepsy | 1 ( 2.3) | 0 ( 0.0) |  |
|  | Hypothyroidism | 1 ( 2.3) | 2 ( 4.8) |  |
|  | malignancy | 1 ( 2.3) | 0 ( 0.0) |  |
|  | HT+IHD | 4 ( 9.3) | 2 ( 4.8) |  |
|  | HT+DM | 6 (14.0) | 5 (11.9) |  |
|  | CVA+HT | 1 ( 2.3) | 1 ( 2.4) |  |
|  | HT+obesity | 2 ( 4.7) | 3 ( 7.1) |  |
|  | HT+DM+IHD | 6 (14.0) | 8 (19.0) |  |
|  | HT+DM+Obesity | 0 ( 0.0) | 2 ( 4.8) |  |
|  | DM+IHD | 1 ( 2.3) | 2 ( 4.8) |  |
|  | DM+Obesity | 0 ( 0.0) | 1 ( 2.4) |  |
| Severity of the disease n (%) | Mild | 25 (33.3) | 25 (33.3) | 1 |
|  | Moderate | 25 (33.3) | 25 (33.3) |  |
|  | Severe | 25 (33.3) | 25 (33.3) |  |

BMI, body mass index; SD, standard deviation; HT, hypertension, RA, rheumatoid arthritis; IHD, ischemic heart disease; DM, diabetes; CVA, cerebrovascular accident, p<0.05 is significant.

### The therapeutic effect of niclosamaide on COVID-19 patients

The current study revealed minor side effects of NCS which were mild-moderate headache lasting for 1-3 days in 76% of patients; mild nausea, vomiting, and diarrhea in 47% of patients lasting 1-2 days; mild abdominal pain for less than one day in 22.7% of patients; temporary skin rash in 18.4% of patients. No major or long-lasting side effects were observed. This indicates the safety of NCS among COVID-19 patients.

The current study did not show any association between death rate of COVID-19 patients and NCS administration (P>0.05) as three patients of severe illness in NCS and in control arms died in this study and as shown in table 2.

**Table 2:**
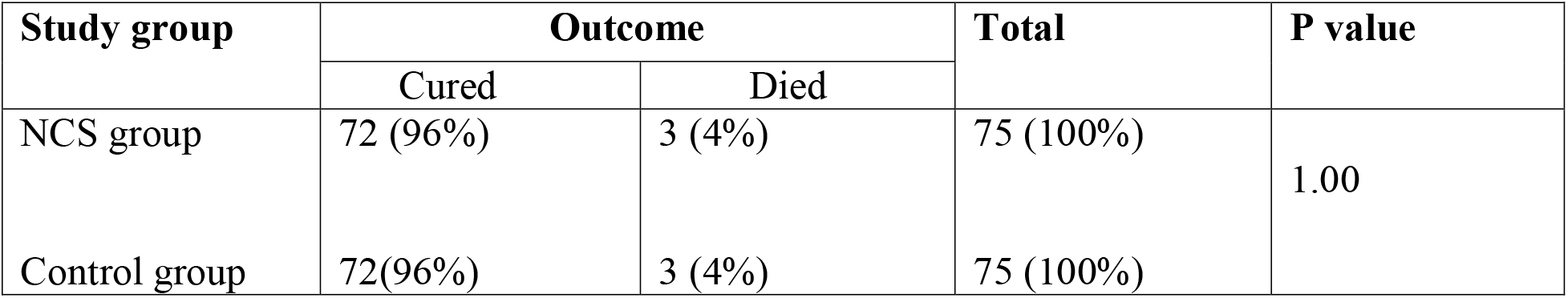
The number and percentage of the cured versus dead COVID-19 patients in NCS compared to control arms.

| Study group | Outcome |  | Total | P value |
| --- | --- | --- | --- | --- |
|  | Cured | Died |  |  |
| NCS group | 72 (96%) | 3 (4%) | 75 (100%) | 1.00 |
| Control group | 72(96%) | 3 (4%) | 75 (100%) |  |

However, it was found that the time to recover after patients being treated in NCS and control groups was significantly three days shorter in NCS, 6.76±4.2 days, than in standard of care control group, 9.76±7.1 days (P=0.007). The accelerated cure course for COVID-19 course was most prominently observed in moderate and severe cases (P<0.05) while mild cases did not show any reduction in time to recover in NCS compare to control group (P>0.05). More to this point, the time to recover after severe patients being treated in NCS and control groups was up to three days shorter in NCS, 8.4±6.3 days, than in standard of care control group, 11.28±8.8 days (P=0.1); nevertheless, the maximal effect of accelerated cure by NCS was found in moderate COVID-19 patients who needed only 6.2±5.2 days to recovery versus 11.8±9.16 days in control group (P-0.01). Thus, moderately ill COVID-19 patients benefited most from adding NCS to therapy by shortening time for recovery more than 5 days and followed by severe patients with 3 days less for recovery as shown in figure 2.

**Figure 2:**
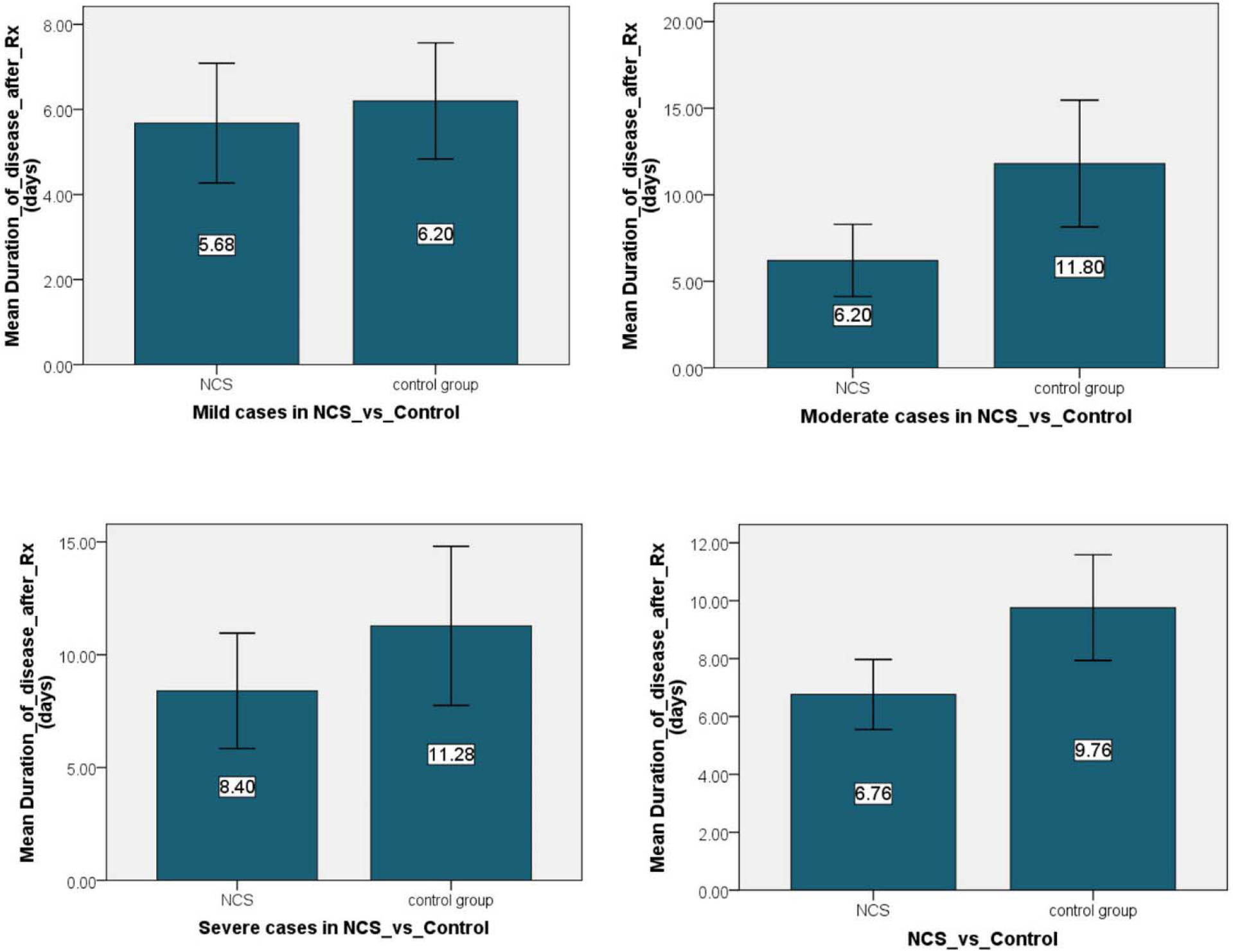
The mean± standard deviation values of the time needed for recovery in mild, moderate and severe groups and in all patients in NCS versus control arms of the study.

It is noteworthy to mention that NCS showed highly observed effect in accelerating the recovery of the disease in those of COVID-19 patients with co-morbidities, about five days less than control group (P<0.0001), than in those patients with no co-morbidities, only one day less than control group (P>0.05), as shown in figure 3.

**Figure 3:**
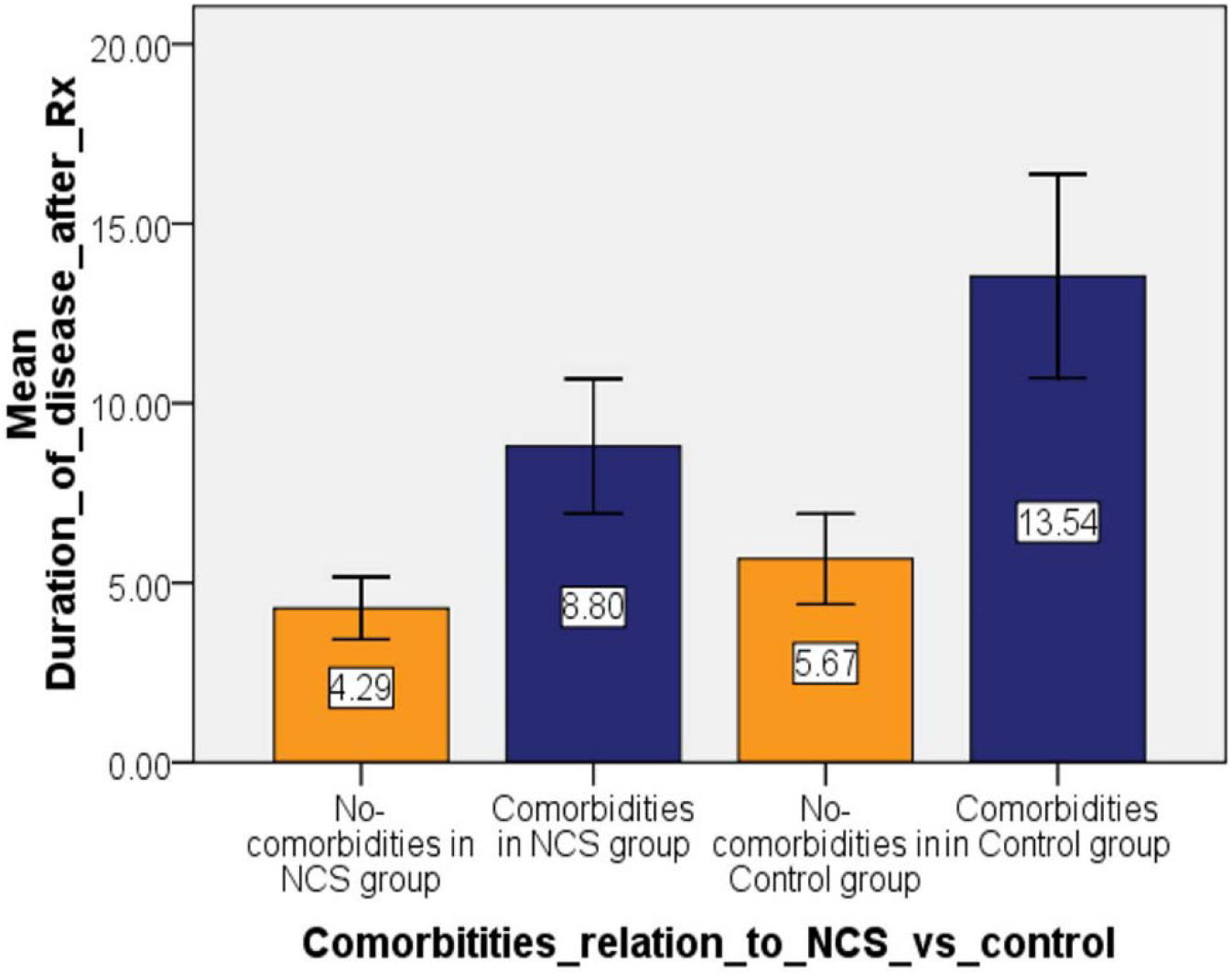
The time needed to recovery for COVID-19 patients with or without co-morbidities in both NCS and control groups.

### Analysis of the secondary attributes of the COVID-19 patients and its effect on recovery

The time to cure after therapy was not different in females, 9±7.1 days, and in males, 7.57±6.5 days (P>0.05). And the six died patients were 50% females and 50% males. On the other hand, the obese patients in this study needed up to two days more time to recovery, 9.8±8.6 days, than non-obese patients, 7.5±5.8 days (P=0.06), as shown in figure 4.

**Figure 4:**
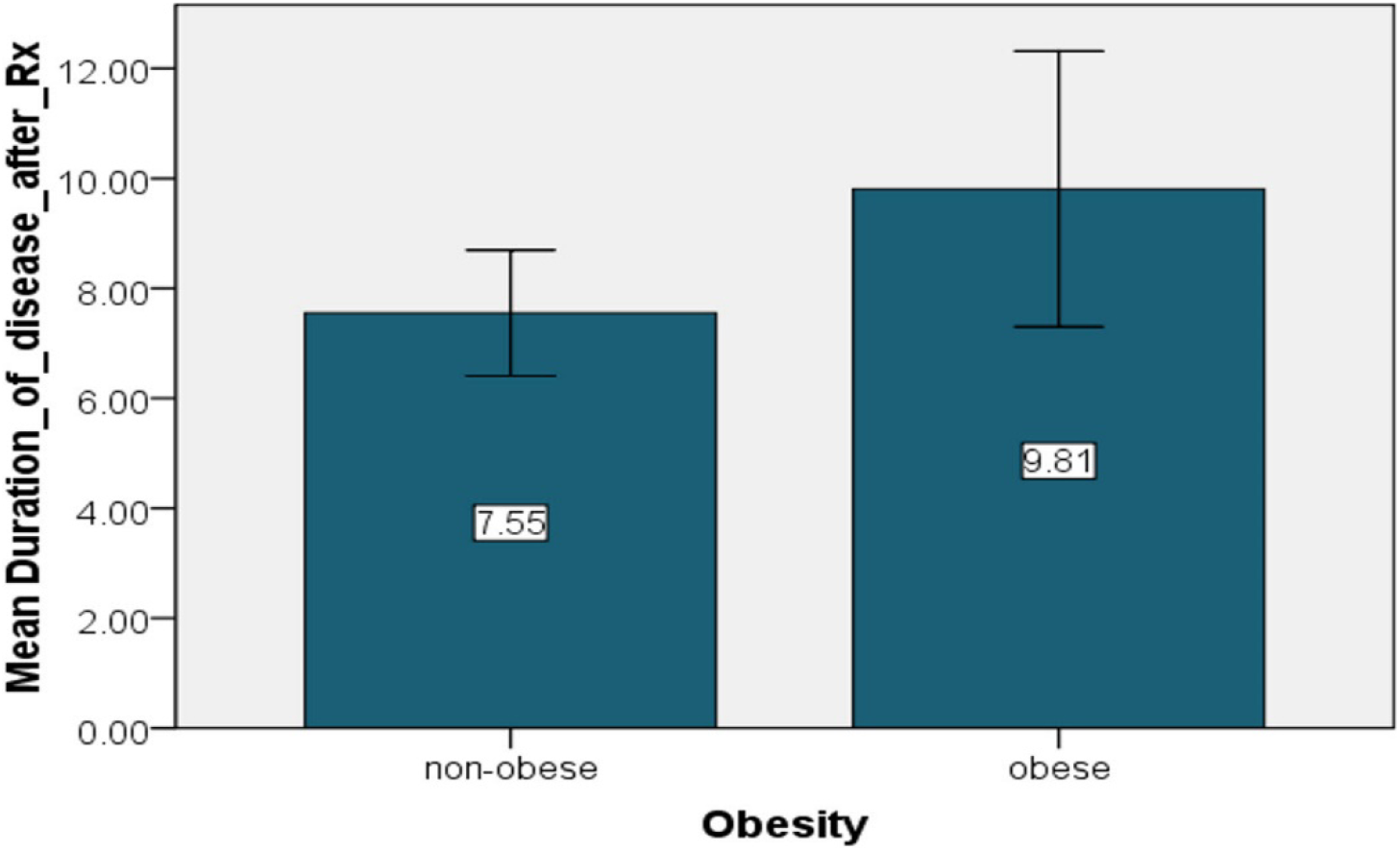
The mean± standard deviation values of the time needed for recovery in obese versus non-obese patients of the study.

In regard to the presence of co-morbidities in COVID-19 patients, the current trial provided evidence that patients with co-morbidities needed six days more to recover, 11.11±7.8 days, than in patients without co-morbidities, 5±3.2 days (P<0.001). In this context, the mean age of patients with co-morbidities was about 17 years higher, 57.13±13.7 year, than of patients with no co-morbidities, 40.35±13.5 year (P<0.001). It is noteworthy to mention that the six patients died were all with co-morbidities (P=0.019). Moreover, patients with co-morbidities showed higher predilection to being moderately to severely ill patients (P<0.0001) as shown in table 3.

**Table 3:** The number and percentage of the mild, moderate, and severe COVID-19 patients in patients with and without co-morbidities

| Comorbidities | Severity of covid 19 |  |  |  | P value |
| --- | --- | --- | --- | --- | --- |
|  | Mild | Moderate | Severe | Total |  |
| Absent | 36(51.4%) | 18 (25.7%) | 16 (22.9%) | 70 (100%) | 0.0001 |
| Present | 14 (17.5%) | 32 (40.0%) | 34 (42.5%) | 80 (100%) |  |

## Discussion

This study was designed to assess the effectiveness and safety of NCS as an addon therapy to the standard measures in COVID-19 management compared to standards of care therapy group.

To our knowledge; it is the first study regarding this aim. For patients with high risk, it is mandatory to search for new or old drugs to be used to lower morbidity and/or mortality of COVID-19 patients. Moreover, COVID-19 patients, especially those who need hospitalization, demand a lot of medical and respiratory supportive care with parenteral fluids, antibiotics, anti-inflammatory and maybe invasive respiratory intubation. Severe/ critical patients may reside more than a month in hospitals. During pandemic, finding drugs that can lower time of hospitalization is important as medical systems all over the world were close to collapse because of huge number of hospitalized COVID-19 patients in respiratory care units. In this endeavor, repurposing old drugs that have potential to lower this burden in SARS-CoV-2 pandemic is necessary. One of the candidates of repurposed drugs to treat COVID-19 patients is NCS.

The current study revealed that NCS did not enhance the survival rate of COVID-19 patients compared to the standard of care control group. Up to 3/75 from NCS and 3/75 patients from control group died and all of them were presented as severe COVID-19 cases. However, NCS use in this study provided evidence that it is useful in reducing significantly the time needed to recovery when compared to control group, about 3 days in severe cases and 5 days in moderate cases while no significant reduction in time to recovery was seen in mild cases. This is the most important finding of this study as 3 to 5 days acceleration of recovery of COVID-19 patients is crucial to intensive care units during pandemic crisis of such fatal and demanding respiratory and multi-organ disease.

Niclosamide could be a candidate for host-directed antiviral therapies. Strategies for controlling viral infections are mainly of two approaches: agents that target the virus directly or agents that target the host [19]. Niclosamide has been reported as a potential agent for host defense during viral infections. It was reported that NCS inhibited SARS-CoV replication and protected Vero E6 cells from cytopathic effects after virus infection [20]. Niclosamide’s effect on anti-viral host defense mechanisms was first reported by Jurgeit et al. They used a monoclonal antibody mabJ2 to stain viral dsRNA in infected cells as a readout for imaged-based screening [21]. They screened a library of 1200 known bioactive compounds and identified NCS as a potent, low micromolar inhibitor of pH-dependent human rhinoviruses (HRV) and influenza virus. The mechanism of action proposed was related to niclosamide’s protonophore activity and its ability to act as a proton carrier [22].

More to the antiviral effect of NCS found in the current study, interestingly NCS succeeded in reducing the time to recovery for COVID-19 patients with co-morbidities, about 5 days, far more than the reduced time to recovery in COVID-19 patients without co-morbidities, only 1 day. Hence, NCS seems beneficial for moderate and severe COVID-19 cases especially those with risk factors and co-morbidities who are most prone to complications and death.

It was reported that NCS as having anti-Chikungunya virus activity through reducing Chikungunya virus entry and transmission [23]. Moreover, NCS was found to be a potent inhibitor of the replication of Zika virus, a mosquito-borne flavivirus, is a growing public health concern following a large outbreak that started in Brazil in 2014 [24].

In addition, repurposing of NCS has been proposed for the treatment of other pulmonary conditions, such as asthma and cystic fibrosis [25]. It has potent bronchodilating effects, and inhibits excessive mucus production [26]. Due to its effects on intracellular Ca2+ levels, NCS also inhibits the release of proinflammatory cytokines such as IL-8, and possibly also other cytokines, which could be of utmost importance to curb the cytokine storm frequently observed in hospitalized Covid-19 patients. Another fortunate aspect is the antibacterial activity of NCS that could be most welcome in fighting potential pulmonary superinfections [27].

This study has some limitations: first, open label study with short time ; second, small sample size and done in single center, However this study, up to the best of our knowledge, was the first study that evaluated NCS effectiveness and safety as add on therapy to the standard care of measures in COVID-19 patients.

In conclusion, this study showed that adding NCS to the standards of care measures had shorter time to stay in the hospital compared with controls especially for patients with moderate and severe COVID-19 who had comorbidities.. In addition, it was relatively safe without observable adverse events. These findings may suggest using NCS as an add on therapy to protocols used for treatment of COVID-19. However, these results are needed to be validated in a larger prospective follow up study.

## Data Availability

The data will be available when needed.

